# Gut microbiota composition correlates with disease severity in myelodysplastic syndrome

**DOI:** 10.1101/2022.04.18.22273768

**Authors:** Giovanna Barbosa Correia Riello, Priscila Mendonça da Silva, Francisca Andrea da Silva Oliveira, Roberta Taiane Germano de Oliveira, Francisco Eliclecio Rodrigues da Silva, Ivo Gabriel da Frota França, Fábio Miyajima, Vânia Maria Maciel Melo, Ronald Feitosa Pinheiro, Danielle S. Macedo

**Affiliations:** Department of Clinical and Toxicological Analysis, School of Pharmacy, Federal University of Ceara, Fortaleza, CE, Brazil; Drug research and Development Center, Faculty of Medicine, Federal University of Ceara, Brazil; Cancer Cytogenomic Laboratory, Drug Research, and Development Center, Department of Clinical Medicine, Faculty of Medicine, Federal University of Ceara, Fortaleza, Brazil; Laboratory of Neuropsychopharmacology, Drug Research and Development Center, Faculty of Medicine, Federal University of Ceara, Fortaleza, CE, Brazil; Laboratory of Microbial Ecology and Biotechnology, Biology Department, Federal University of Ceará, Fortaleza, Brazil; National Institute for Translational Medicine (INCT-TM, CNPq), Ribeirão Preto, SP; Oswaldo Cruz Foundation (Fiocruz), Branch Ceara, Eusebio, Brazil

**Keywords:** gut microbiota composition, Myelodysplastic Syndromes, 16S rRNA gene, high-risk MDS, elderly people

## Abstract

The myelodysplastic syndrome (MDS) is a heterogeneous group of clonal disorders of hematopoietic progenitor cells related to ineffective hematopoiesis and an increased risk of transformation to acute myelogenous leukemia. MDS is divided into categories, namely lineage dysplasia (MDS-SLD), MDS with ring sideroblasts (MDS-RS), MDS with multilineage dysplasia (MDS-MLD), MDS with excess blasts (MDS-EB). The International Prognostic Classification System (IPSS) ranks the patients as very low, low, intermediate, high, and very high based on disease evolution and survival rates. Evidence points to toll-like receptor (TLR) abnormal signaling as an underlying mechanism of this disease, providing a link between MDS and immune dysfunction. Notably, microbial signals, such as lipopolysaccharide from gram-negative bacteria can activate or suppress TLRs. Therefore, we hypothesized that MDS patients present gut microbiota alterations associated with disease subtypes and prognosis. We sequenced the 16S rRNA gene from fecal samples of 30 MDS patients and 16 healthy elderly controls to test this hypothesis. We observed a negative correlation between Prevotella spp. and Akkermansia spp. in MDS patients compared with the control group. High-risk patients presented a significant increase in the genus Prevotella spp. in relation to the other risk categories. There was a significant reduction in the abundance of the genus Akkermansia spp. in high-risk patients compared with low- and intermediate-risk. There was a significant decrease in the genus Ruminococcus spp. in MDS-EB patients compared with controls. Our findings show a new association between gut dysbiosis and higher-risk MDS, with a predominance of gram-negative bacteria in the gut of these patients.

## 1. Introduction

The myelodysplastic syndrome (MDS) is a heterogeneous group of clonal disorders of hematopoietic progenitor cells characterized by cytopenia, dysplasia due to ineffective hematopoiesis, and associated increased risk of transformation to acute myeloid leukemia (AML) [1]. As most patients with MDS are elderly (median age range 65 to 70 years), its incidence and prevalence are rising as the population ages [2].

Based on the risk of progression to AML, MDS is divided into high-and low-risk categories. Cytogenetic and molecular abnormalities, hemoglobin level, platelet and neutrophil counts, transfusion dependency, and percentage of blasts in the bone marrow are variables used to distinguish between high and low MDS risk [3,4]. Most importantly, the Revised International Prognostic Scoring System (R-IPSS), published by Greenberg et al in 2012 [4], is still the more robust way to predict the overall survival and risk of AML transformation.

The pathogenesis of MDS involves RNA splicing, epigenetic regulation DNA repair system, mitotic and spindle genes, and chronic immune stimulation [5]. Chromosomal abnormalities are present in approximately 50% of de novo MDS, whereas somatic point mutations can be identified in up to 95% of cases [6]. Furthermore, an inflammatory microenvironment is present in MDS. This disease is characterized by immune dysfunction, stromal microenvironment, and cytokine imbalance [7].

To our knowledge, no previous research was conducted to assess the role of the host gut microbiota in MDS. However, it is worth noting that earlier research has suggested that the gut microbiota plays a role in hematopoiesis. Indeed, the gut microbiota appears to have an impact on iron metabolism and hepcidin induction [8], platelet disorders [9], and post-transplant reactions [10]. Furthermore, besides hematopoiesis, gut microbiota also regulates inflammation by influencing the differentiation of inflammatory cell types and cytokine production [11]. Hence, the knowledge about gut microbiota modifications in MDS patients will open new avenues for understanding the mechanisms underpinning this syndrome and developing novel therapeutic strategies, such as probiotics.

We hypothesized that MDS patients present gut microbiota alterations associated with disease subtypes in the present study. Therefore, our primary outcome was to identify and analyze the association between gut microbiota diversity in patients with MDS compared with the gut microbiota of the elderly without hematological disorders. Our secondary outcome was to investigate the association between gut microbiota profile in MDS patients and disease prognosis.

## 2. Materials and Methods

### 2.1 Patients and Ethical Aspects

The institutional review board of the Federal University of Ceara/PROPESQ-UFC (number: 58761816.2.0000.5054) approved the study. All participants read and signed the written informed consent. Furthermore, the study complied with the ethical precepts of the research, based on Resolution 466/12 of Brazil’s National Health Council and the Declaration of Helsinki.

This study included patients diagnosed with MDS aged 45 to 95 years who attended the Drug Research and Development Center (NPDM) of the Federal University of Ceara. MDS patients (n=27) were classified according to 2016 WHO classification. Twenty-one patients were classified as early MDS (MDS-SLD, MDS-RS, MDS-MLD) and six as advanced MDS (MDS-EB1/EB2). Patients were also classified according to the R-IPSS score as low (very low + low, n=17); intermediate (n=5) and high risk (high + very high, n=5) (Table 1). The control group comprised 16 healthy elderly individuals. The exclusion criteria were patients with heart disease, pneumopathy, liver disease, history of alcoholism, psychiatric disorders, and those on chemotherapy or had used antibiotics 30 days before the samples collection.

**Table 1.**
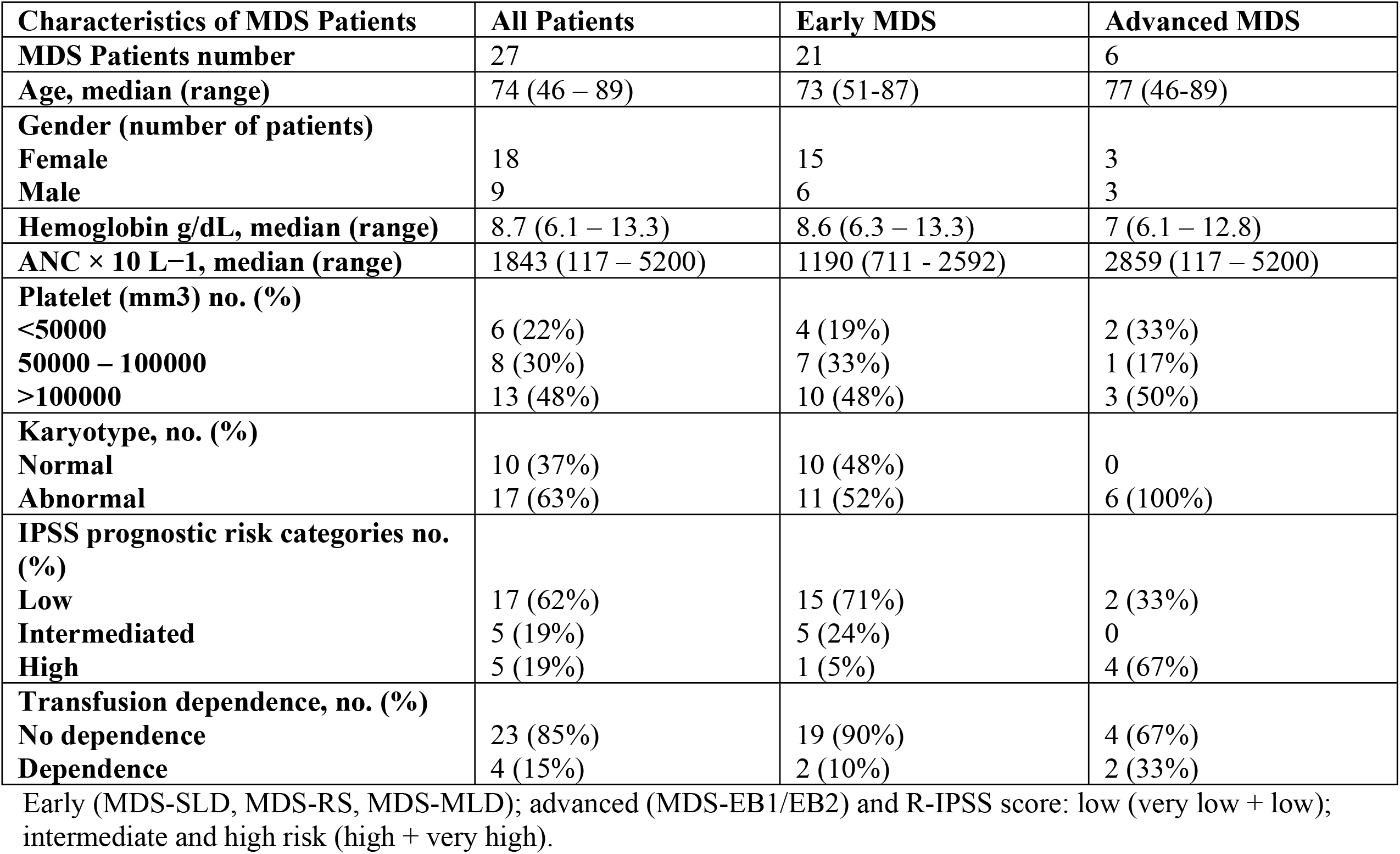
Clinical characteristics of MDS patients

| Characteristics of MDS Patients | All Patients | Early MDS | Advanced MDS |
| --- | --- | --- | --- |
| **MDS Patients number** | 27 | 21 | 6 |
| **Age, median (range)** | 74 (46 – 89) | 73 (51-87) | 77 (46-89) |
| **Gender (number of patients)** |  |  |  |
| Female | 18 | 15 | 3 |
| Male | 9 | 6 | 3 |
| **Hemoglobin g/dL, median (range)** | 8.7 (6.1 – 13.3) | 8.6 (6.3 – 13.3) | 7 (6.1 – 12.8) |
| **ANC × 10 L<sup>-1</sup>, median (range)** | 1843 (117 – 5200) | 1190 (711 - 2592) | 2859 (117 – 5200) |
| **Platelet (mm<sup>3</sup>) no. (%)** |  |  |  |
| <50000 | 6 (22%) | 4 (19%) | 2 (33%) |
| 50000 – 100000 | 8 (30%) | 7 (33%) | 1 (17%) |
| >100000 | 13 (48%) | 10 (48%) | 3 (50%) |
| **Karyotype, no. (%)** |  |  |  |
| Normal | 10 (37%) | 10 (48%) | 0 |
| Abnormal | 17 (63%) | 11 (52%) | 6 (100%) |
| **IPSS prognostic risk categories no. (%)** |  |  |  |
| Low | 17 (62%) | 15 (71%) | 2 (33%) |
| Intermediated | 5 (19%) | 5 (24%) | 0 |
| High | 5 (19%) | 1 (5%) | 4 (67%) |
| **Transfusion dependence, no. (%)** |  |  |  |
| No dependence | 23 (85%) | 19 (90%) | 4 (67%) |
| Dependence | 4 (15%) | 2 (10%) | 2 (33%) |

Early (MDS-SLD, MDS-RS, MDS-MLD); advanced (MDS-EB1/EB2) and R-IPSS score: low (very low + low); intermediate and high risk (high + very high).

Before sample collection, the patients answered a questionnaire with their personal information, eating habits, possible medication use, intestinal symptoms, and pre-existing diseases. See questionnaire in Supplementary Methods

### 2.2 Sample collection and preparation

The stool samples were collected on the day of the patient’s consultation. The samples were transported to the laboratory within an icebox (not exceeding four hours) and aliquoted in cryotubes. The samples were kept at −80 °C until the DNA extraction was performed.

### 2.3 DNA extraction

Firstly, the samples were vigorously homogenized using a MiniBeadBeater (BioSpec, Bartlesville, OK, USA) to effectively liberate bacterial DNA from complex fecal biomass (THOMAS, 2015). For this homogenization, we used samples around 0.3 g, 0.1 mm glass beads (BioSpec, Bartlesville, OK, USA) incubated at 95 °C in lysis buffer (QIAamp® Fast DNA Stool Mini Kit, QIAGEN Inc., Valencia, CA, USA).

After that, the DNA was extracted and purified from 0.2 g (total lysed humid weight) of stool using the QIAamp® Fast DNA Stool Mini Kit, according to the manufacturer’s instructions.

We evaluated the quality of extracted DNA with the Nanodrop ND-1000 spectrophotometer (Thermo Scientific, Waltham, MA, USA) and confirmed by electrophoresis in 0.8% agarose gel with 1 × TAE buffer. In addition, the DNA was quantified by Qubit® 2.0 fluorometer using the dsDNA BR Assay kit (Invitrogen(tm), SP, Brazil). Extracted DNA was stored at −20 °C until analysis.

### 2.4 Sequence processing and data analysis

We generated the amplicon library using primers (515F / 806R) targeting the V4 region of the 16S rDNA. First, amplification was performed using 25 μL of a reaction containing the following reagents: 14.8 μL of nuclease-free water (Certified Nuclease-free, Promega, Madison, WI, EUA), 2.5 μL of 10x high-fidelity reaction buffer (Invitrogen, Carlsbad, CA, USA), 1.0 μL of 50 mM MgSO4, 0.5 μL of each primer (10 μM concentration, 200 pM final concentration), 1.0 unit of Taq Polymerase High Fidelity Platinum (Invitrogen, Carlsbad, CA, USA) and 4.0 μL of DNA (10 ng). The conditions for PCR were as follows: 94 °C for 4 min to denature the DNA, with 35 cycles at 94 °C for 45 s, 50 °C for 60 s, and 72 °C for 2 min, with a final extension of 10 min at 72 °C. In addition, a specific Illumina Nextera XT index pair (Illumina, San Diego, CA) was added to the purified PCR product for indexing. Thus, each 50 μL reaction contained: 23.5 μL of nuclease-free water (Certified Nuclease-free, Promega, Madison, WI, USA), 5.0 μL of 10× High Fidelity PCR Buffer (Invitrogen, Carlsbad, CA, USA), 4.8 μL of 25 mM MgSO4, 1.5 μL of dNTP (10 mM each), 5.0 μL of each Nextera XT index (Illumina, San Diego, CA, USA), 1.0 unit of Platinum Taq polymerase High Fidelity (Invitrogen, Carlsbad, CA, USA), and 5.0 μL of each purified PCR product.

The conditions for this second round PCR were as follows: 95 °C for 3 min to denature the DNA, with 12 cycles at 95 °C for 20 s, 55 °C for 30 s, and 72 °C for 30 min, with a final extension of 5 min at 72 °C.

After adding the index, the PCR products were cleaned, as previously described, and quantified using the dsDNA BR assay kit (Invitrogen, Carlsbad, CA, USA) on a Qubit 2.0 fluorometer (Invitrogen, Carlsbad, CA, USA). Once quantified, the DNA samples were normalized to leave them in an equimolar concentration (16 nM). In parallel, the positive control Phix (Illumina, San Diego, CA, USA) was prepared, diluting it to 12 pM and denatured in the same way as the sample pool. Next, the molarity of the pool was determined by fluorometric quantitation, diluted to 4 nM, denatured with 0.2N NaOH, and then diluted to a final concentration of 10 pM together with 20% PhiX for loading in the MiSeq sequencer (Illumina, San Diego, CA, USA).

Illumina adapter sequences were trimmed from the already demultiplexed raw fastq files using Cutadapt v1.8 in paired-end mode. The quality of the reads was assessed using FastQC v.0.11.8 (Martin, 2013) and vsearch v2.10.4 (Rognes, Flouri, Nichols, Quince, & Mahé, 2016). Subsequent analyses were performed within the R v3.5.3 environment (R Development Core Team, 2016), following DADA2 v1.11.1 (B. J. Callahan et al., 2016) package authors suggested pipeline and adjusting parameters to our data. The product was non-chimeric amplicon sequence variants (ASVs) (B. J. Callahan, Mcmurdie, & Holmes, 2017) table, which records the number of times each ASV (sequence differing by as little as one nucleotide) was observed in each sample. Taxonomy assignment of representative ASVs was performed with DADA2 against SILVA 132 reference database (B. Callahan, 2018). Finally, we performed downstream analyses using the phyloseq v1.26.1 package (Mcmurdie & Holmes, 2013) and graphical representations with ggplot2 v3.1(Wickham, 2009).

### 2.5 Analysis of α-diversity and species richness

Alpha-diversity estimators were calculated and tested for normality by the Shapiro-Wilk test. As the Shannon diversity and Simpson evenness were parametric, a one-way analysis of variance and Tukey’s honestly significant difference (HSD) post hoc tests were used for multiple comparisons of means at a 95% confidence interval. For richness, Chao1 index, and Observed ASVs, we used the Kruskal-Wallis non-parametric test.

### 2.6 Statistical Analysis

Initially, the BIOENV tool (part of the vegan package for R) was used to select the subset of variables that best explain the variance of biological data, using biotic dissimilarity matrices and the data for each patient. Then, this subset of variables, defined for each sample, was used in the PERMANOVA test (adonis2 function of the vegan package), a non-parametric method like the analysis of variance. PERMANOVA test uses permutation methods to test differences between groups to define their significance and correlation with biotic data [12]. Finally, P-value was set at P< 0.05.

## 3 Results

### 3.1 Clinical characteristics, α-diversity, and the richness of gut microbiota in MDS patients

Table 1 presents the characteristics, WHO classification, and R-IPSS score of the patients included in this study. Based on the analysis of α-diversity and richness of the samples, no differences were observed in the Shannon, Simpson, and Chao1 indices, comparing the median values obtained in the control and MDS groups. This indicates that the α-diversity and the richness of the samples are equivalents between the two groups studied (Fig. 2).

**Figure 1.**
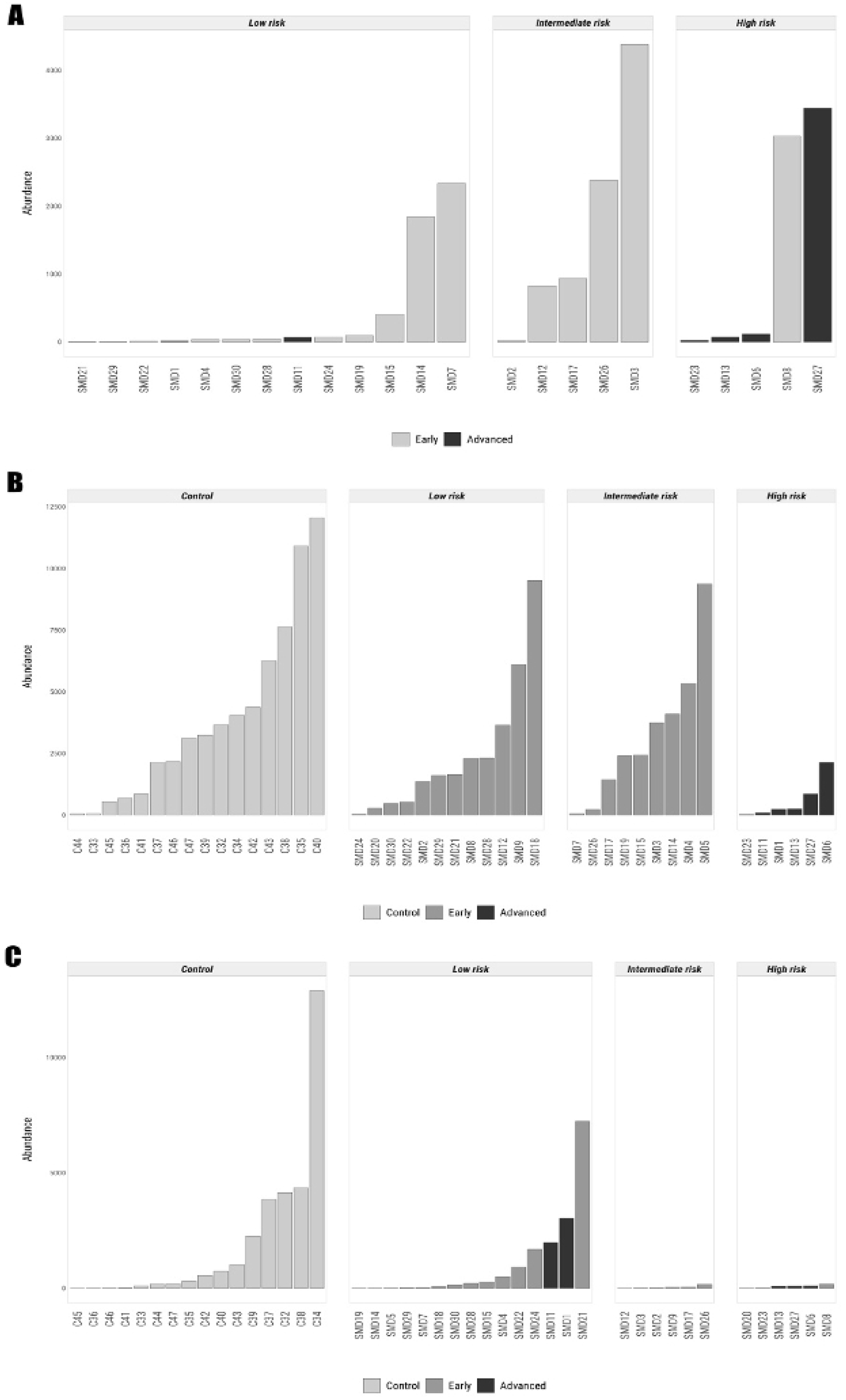
Distribution of the genus *Prevotella* (A), family Ruminococcaceae (B), and genus *Akkermansi*a (C) according to the revised International Prognostic Scoring System (R-IPSS) prognostic score in MDS patients and control samples. Patients are identified according to their MDS category.

**Figure 2.**
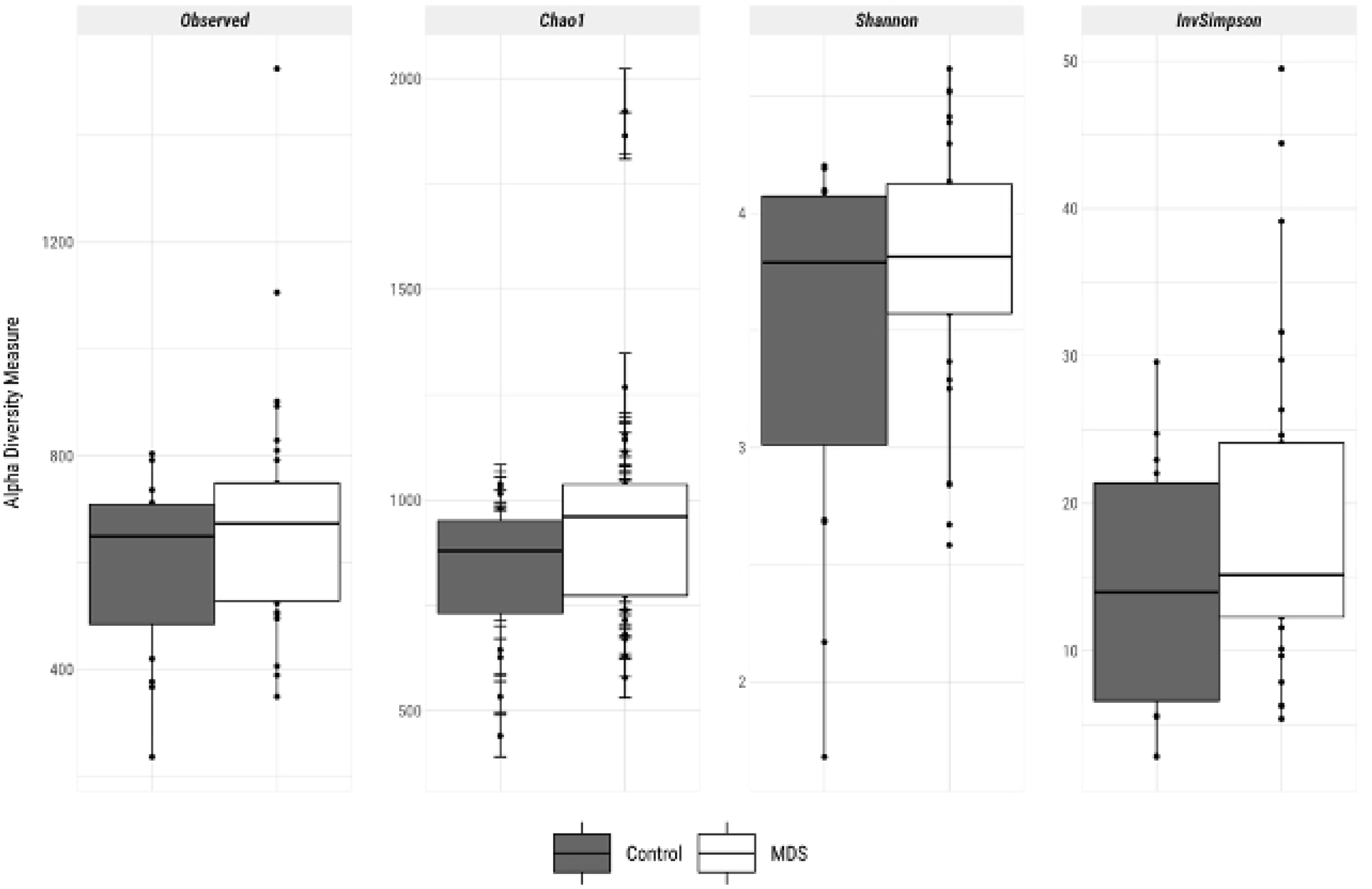
α-diversity and the richness of the samples between control and MDS groups.

Based on the Spearman correlation test, there was a negative correlation (−0.42) between *Prevotella* spp. And *Akkermansia* spp. in MDS patients when compared with the control group (Fig. 3)

**Figure 3.**
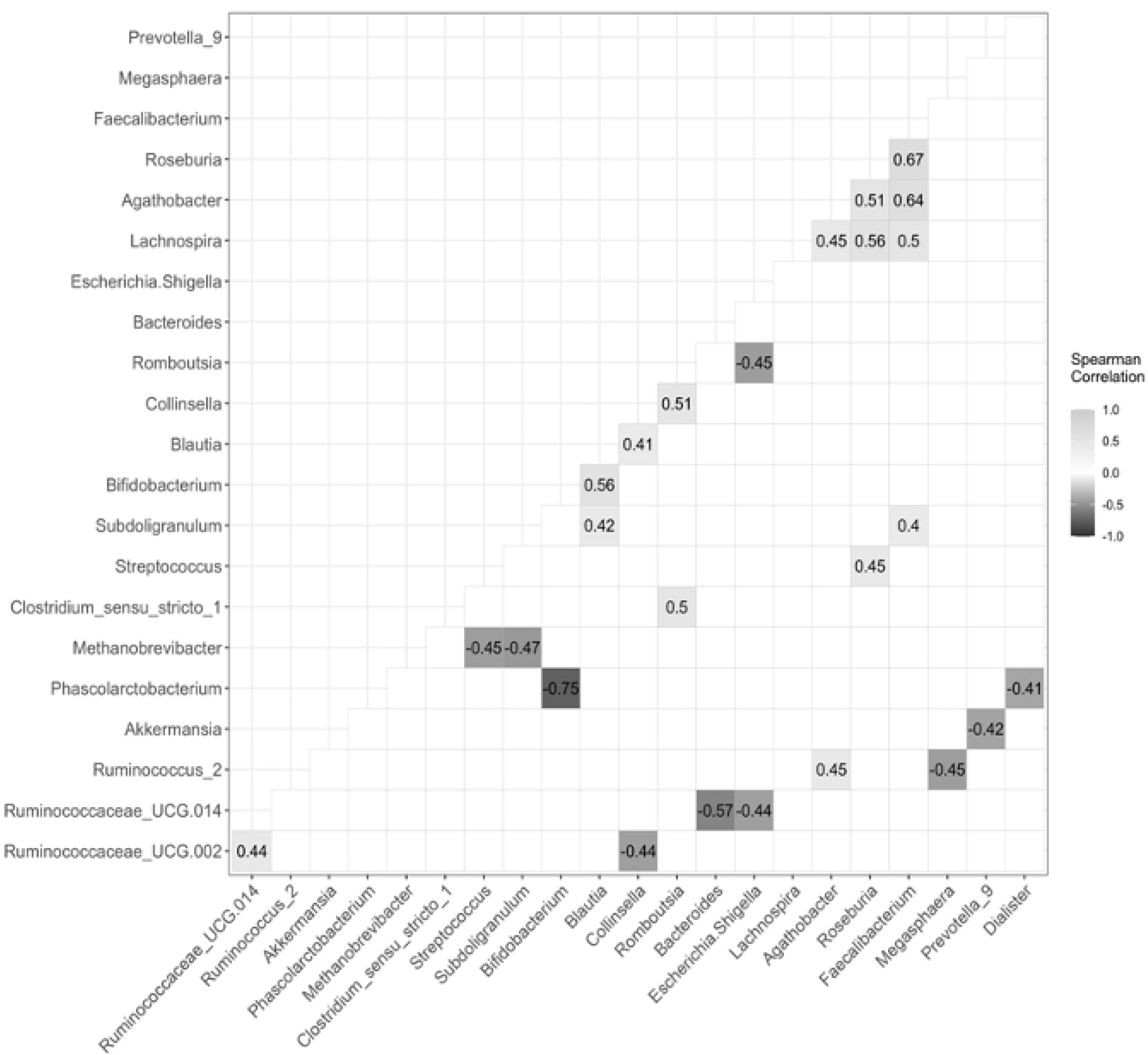
Correlations between the composition of the fecal microbiota of control and MDS groups based on Spearman correlation test.

### 3.2 High-risk patients based on R-IPSS have a unique profile: increase of Prevotella and decrease in Akkermansia

High-risk patients presented a significant increase in the genus *Prevotella* spp. in relation to the other risk categories (P= 0.011) (Fig. 1A). On the other hand, there was a significant reduction in the abundance of the genus *Akkermansia* in the group of high-risk patients in comparison with low- and intermediate-risk (P = 0.005) (Fig. 1C).

### 3.3 MDS subtypes according to WHO 2016

Regarding the genus distribution, there was a significant decrease in the genus *Ruminococcus* in MDS-EB1/EB2 patients compared with controls (P= 0.004) (Fig. 1B).

## 4 Discussion

For the first time, as far as we know, we are showing the presence of gut microbiota alterations in MDS patients. Furthermore, high-risk patients presented the main alterations, providing novel mechanisms underpinning this syndrome severity.

A wide range of inflammatory disorders presents with changes in gut microbiota composition, as observed in human and animal studies [13]. Furthermore, gut microbiota influences hematopoiesis [14].

Our results revealed that high-risk MDS patients presented a significant increase in the genus *Prevotella*. Notably, among the vast composition of the human intestinal microbiota, the exacerbated abundance of the genus *Prevotella*, known as a biomarker of one of the three proposed human intestinal enterotypes [15], has been associated with the manifestation of inflammatory diseases, such as rheumatoid arthritis [16], and inflammatory bowel disease [17]. Our first defense against infection is initiated by innate immunity, and the recognition of pathogens occurs by pattern recognition receptors (PRRs) expressed in hematopoietic and no hematopoietic cells. Under normal conditions, hematopoietic cells (HSC) are in a quiescent state. At the same time, in response to inflammation, they proliferate and signal through toll-like receptors (TLRs). The sustained exposure to TLR signals is associated with loss of normal HSC function, negatively influencing these cells towards apoptosis and myeloid differentiation. This mechanism has been an important phenomenon in bone marrow disorders such as myelofibrosis, acute myeloid leukemia, and MDS [18].

*Prevotella* genus, more precisely the *P. intestinalis* species, promotes inflammation and intestinal dysbiosis, an effect that is shared with other intestinal bacteria. These damaging effects of *Prevotella* on the host’s physiology can be caused by several factors, including direct effects of this bacteria or mediated by modification of intestinal microbiome composition. Thus, the abundance of this genus is interpreted as a marker of a new microbiome pattern, which is potentially inflammatory. Furthermore, the increase in the severity of the intestinal inflammatory process is associated with changes in the microbial ecosystem and the metabolites produced in this microenvironment [19].

Previous studies have shown that inflamed tissues of mice colonized with *P. intestinalis* present increased levels of interleukin (IL)-6 and tumor necrosis factor (TNF)-α, in addition to higher levels of pro-inflammatory chemokines, with consequent neutrophil infiltration. It is also well demonstrated that colonization by *Prevotella* shapes the host’s immunity, causing a reduction in IL-18 production. Furthermore, previous studies have shown that reducing IL-18 production by 1.3-1.5 times is sufficient to disturb the homeostasis of the intestinal microenvironment [20]. MDS has been constantly reported with increased IL-6 and TNF-α, which also may predispose to apoptosis, a critical phenomenon related to anemia in MDS. These cytokines create bone marrow with modifications called the inflammasome. The prototypical example of a hematological disease caused by bone marrow inflammation is aplastic anemia, similar to hypoplastic MDS [21].

*Prevotella* spp. also influences the production of short-chain fatty acids (SCFAs). SCFAs are beneficial for intestinal health since they are a source of energy for cells and contribute to maintaining intestinal barrier function, being indicated even as protectors against the development of colorectal cancer and in the control of local inflammatory processes [22]. The intestinal production of SCFAs is influenced by the composition of the diet and the microbiota [23]. Hence, there is an inverse relationship between the prevalence of *P. intestinalis* and the levels of SCFAs produced in the intestine [19]. Furthermore, Iljazovic 2020 and collaborators [19] showed that the intestinal colonization of *Prevotella* sp. in mice causes a decrease in the relative abundance of the phyla Bacteroidetes and Firmicutes (Lachnospiraceae and Ruminococcaceae), which may explain the reduction in the production of SCFAs. Notably, the Ruminococcaceae is an important family of bacteria producing SCFAs [24]. Regarding MDS, we know that DNA methylation is prevalent in tumor suppressor genes, principally MDS-EB1/EB2. Thus, SCAFs may modify epigenetics, possibly influencing the hypomethylating agents commonly used to treat MDS-EB1/EB2, such as azacytidine and decitabine. In addition, SCAFs can also act as histone deacetylase (HDACs) inhibitors, changing DNA structure and transcriptions [25].

It must be emphasized that the reduction in the genus *Ruminococcus*, observed here in advanced MDS patients, i.e., MDS-EB1/EB2 subtype, may increase the inflammatory processes that have been constantly reported in bone marrow disorders. We do not know if this effect is related to reducing SCFAs, but this result creates a new idea that deserves further evaluation.

Given this well-known set of changes promoted by the predominance of the *Prevotella* in the intestinal microenvironment, these changes can affect the intestinal barrier homeostasis [19]. Indeed, changes in the intestinal barrier contribute to bacterial translocation and metabolites into the individual’s bloodstream. Based on our findings, we can speculate that possible changes in the intestinal barrier caused by *Prevotella* in MDS patients may be related to alterations in IL-18 since this cytokine has a protective role in preventing dysbiosis [26], promoting the integrity and regeneration of the epithelial barrier [27]. As far as we know, to date, there is no study evaluating intestinal barrier permeability in MDS patients. Despite this, MDS patients present a higher expression of the CD40 receptor on monocytes, contributing to bone marrow failure through CD40–CD40L interactions with T helper cells [28]. Since lipopolysaccharides (LPS) are regulators of CD40 expression in macrophages [29], we can infer that the gut dysbiosis detected here in MDS patients is causing bacterial translocation and influencing the peripheral immune response these patients by LPS release to the plasma. Therefore, intestinal permeability, IL-18, and LPS plasma levels in MDS patients must be evaluated in future studies since this mechanism may contribute to the inflammatory alterations observed in these patients.

Notably, TLR-4, which mediates the recognition of LPS, is the best-described receptor in MDS. The constant activation of TLR-4 is associated with DNA damage induced by reactive oxygen species (ROS), generating the accumulation of genotoxic components [30], and, consequently, chromosomal abnormality [31]. Indeed, LPS activation of TLR-4 receptors may justify some inflammatory alterations observed in MDS, such as the NLRP3-mediated inflammasome activation that triggers the permanent release of pro-inflammatory cytokines such as IL-1β [32].

Additionally, increased plasma LPS activates the nucleotide-binding and oligomerization domain (NOD)-1 pathway in stromal cells, acting in the hematopoiesis process. The TLR and NOD1 pathways share similar signaling molecules, such as factor-3 associated with the TNF receptor (TRAF3), which signals interferon production, activating the JAK-STAT signaling pathway, activating a genetic profile necessary to promote hematopoiesis [14]. Thus, we can assume that an imbalance in the composition of the microbiota and the maintenance of the intestinal barrier may result in changes in the hematopoiesis process, which would be even more severe for patients with MDS, which already presents alterations in hematopoiesis.

Still concerning the intestinal barrier, interestingly, our study found a negative correlation between the genera *Prevotella* and *Akkermansia* in SMD MDS patients. The *A. muciniphila* controls the mucus barrier in the intestinal epithelium that functions as a protective physical barrier by acting against toxic metabolites present in the intestine and controlling translocation events. Therefore, this species acts by degrading mucin when it is in excess, and stimulating its production by goblet cells of the intestinal epithelium, when it is scarce [33]. Of utmost importance, the use of *A. muciniphila* as microbial therapy for altering immunotherapy response was reported by Routy et al. [34], who demonstrated reversion of rapid tumor growth and antibiotic-induced dysbiosis using anti-programmed death 1 immunotherapy plus *A. muciniphila*. Although still rare, this type of therapy is growing, and its evidence increase day by day [34].

The present study has some limitations since we did not perform experiments to detect plasma LPS, IL-18 alterations, and SCFAs levels, which would help us explain the mechanisms suggested here.

In conclusion, the present study’s findings show a new association between dysbiosis (with the predominance of gram-negative bacteria) and higher-risk MDS. This dysbiosis can contribute to a systemic inflammatory profile in these patients, hindering the clinical management and a favorable outcome for the patient’s survival. Thus, we can propose that intestinal dysbiosis, observed in higher-risk MDS patients, can contribute to LPS translocation to the bloodstream leading to TLR-4 and NLRP3 activation. This activation results in chronic inflammatory alterations observed in MDS patients, promoting DNA damage and chromosomal abnormalities. This study opens new avenues for studying the influence of gut microbiota composition in response to hypomethylating agents in MDS.

## Data Availability

The data underlying the results presented in the study are available from the author Prof. Giovanna Riello.

## Acknowledgment

The authors thank the Genomics and Bioinformatics Central (CEGENBIO) of the Drug Research and Development Center of Federal University of Ceara for technical support. In addition, the authors acknowledge the Brazilian Institutions, CNPq, CAPES, and FUNCAP for the financial support of the study.

## Declarations Ethics approval

The institutional review board of the Federal University of Ceara/PROPESQ-UFC (number: 58761816.2.0000.5054) approved the study. All participants read and signed the written informed consent. Furthermore, the study complied with the ethical precepts of the research, based on Resolution 466/12 of Brazil’s National Health Council and Declaration of Helsinki.

## Consent to participate

Written informed consent was obtained from each participant and is available under request directly to.

## Author Contributions

GBCR, VMMM, RFP, DSM – conceptualization, design of methods, writing - Original Draft.

GBCR, RFP, RTGO, IGFF – Investigation.

PMS, FASO, IGFF – Validation.

RTGO, FERS, FASO, FM, VMMM – Formal analysis.

GBCR, DSM, RFP - Writing - Review & Editing

VMMM - Funding acquisition

GBCR - Data Curation

All authors contributed to the final version of the MS.

## Availability of data and material

16S data is available under request directly to Prof. Giovanna Riello.

## Consent for publication

The patients gave their written informed consent for publication of their clinical details. A copy of the consent is available for review.

## References

1. Jonas BA, Greenberg PL. MDS prognostic scoring systems - Past, present, and future. Best Practice and Research: Clinical Haematology. Bailliere Tindall Ltd; 2015. pp. 3– 13. doi:10.1016/j.beha.2014.11.001

2. Ria R, Moschetta M, Reale a., Mangialardi G, Castrovilli a., Vacca a., et al. Managing myelodysplastic symptoms in elderly patients. Clinical interventions in aging. 2009;4: 413–423. doi:10.2147/CIA.S5203

3. Bejar R, Abdel-Wahab O. The importance of subclonal genetic events in MDS. Blood. 2013;122: 3550–3551. doi:10.1182/blood-2013-09-527655

4. Greenberg P, Tuechler H. Revised international prognostic scoring system for myelodysplastic syndromes. …. 2012;120: 2454–2465. doi:10.1182/blood-2012-03-420489.The

5. Lindsley RC, Ebert BL. Molecular pathophysiology of myelodysplastic syndromes. Annual review of pathology. 2012/08/28. 2013;8: 21–47. doi:10.1146/annurev-pathol-011811-132436

6. Bernard E, Nannya Y, Hasserjian RP, Devlin SM, Tuechler H, Medina-Martinez JS, et al. Implications of TP53 allelic state for genome stability, clinical presentation and outcomes in myelodysplastic syndromes. Nature Medicine. 2020;26: 1549–1556. doi:10.1038/s41591-020-1008-z

7. Yang L, Qian Y, Eksioglu E, Epling-Burnette PK, Wei S. The inflammatory microenvironment in MDS. Cellular and Molecular Life Sciences. 2015;72: 1959– 1966. doi:10.1007/s00018-015-1846-x

8. Peyssonnaux C, Zinkernagel AS, Datta V, Lauth X, Johnson RS, Nizet V. TLR4-dependent hepcidin expression by myeloid cells in response to bacterial pathogens. Blood. 2006;107: 3727–3732. doi:10.1182/blood-2005-06-2259

9. Cines DB, Liebman H, Stasi R. Pathobiology of Secondary Immune Thrombocytopenia. Seminars in Hematology. 2009;46. doi:10.1053/j.seminhematol.2008.12.005

10. Taur Y, Jenq RR, Perales MA, Littmann ER, Morjaria S, Ling L, et al. The effects of intestinal tract bacterial diversity on mortality following allogeneic hematopoietic stem cell transplantation. Blood. 2014;124: 1174–1182. doi:10.1182/blood-2014-02-554725

11. Brandsma E, Kloosterhuis NJ, Koster M, Dekker DC, Gijbels MJJ, van der Velden S, et al. A Proinflammatory Gut Microbiota Increases Systemic Inflammation and Accelerates Atherosclerosis. Circulation Research. 2019;124: 94–100. doi:10.1161/CIRCRESAHA.118.313234

12. Anderson M, Gorley RN, Clarke KR. PERMANOVA + for PRIMER user manual. 2008;1: 1:218. doi:10.1016/j.isatra.2014.07.008

13. Blander JM, Longman RS, Iliev ID, Sonnenberg GF, Artis D. Regulation of inflammation by microbiota interactions with the host. Nature Immunology. 2017;18: 851–860. doi:10.1038/ni.3780

14. Yan H, Baldridge MT, King KY. Hematopoiesis and the bacterial microbiome. Blood. 2018;132: 559–564. doi:10.1182/blood-2018-02-832519

15. Arumugam M, Raes J, Pelletier E, Le Paslier D, Yamada T, Mende DR, et al. Enterotypes of the human gut microbiome. Nature. 2011;473: 174. doi:10.1038/nature09944

16. Scher JU, Sczesnak A, Longman RS, Segata N, Ubeda C, Bielski C, et al. Expansion of intestinal Prevotella copri correlates with enhanced susceptibility to arthritis. eLife. 2013;2: 1–20. doi:10.7554/elife.01202

17. Su T, Liu R, Lee A, Long Y, Du L, Lai S, et al. Altered intestinal microbiota with increased abundance of prevotella is associated with high risk of diarrhea-predominant irritable bowel syndrome. Gastroenterology Research and Practice. 2018;2018: 11–13. doi:10.1155/2018/6961783

18. Leimkühler NB, Schneider RK. Inflammatory bone marrow microenvironment. Hematology (United States). 2019;2019: 294–302. doi:10.1182/hematology.2019000045

19. Iljazovic A, Roy U, Gálvez EJC, Lesker TR, Zhao B, Gronow A, et al. Perturbation of the gut microbiome by Prevotella spp. enhances host susceptibility to mucosal inflammation. Mucosal Immunology. 2020. doi:10.1038/s41385-020-0296-4

20. Maayan Levy, Thaiss CA, Zeevi D, Dohnalová1 L, Zilberman-Schapira G, Mahdi JA, et al. 乳鼠心肌提取 HHS Public Access. Physiology& behavior. 2017;176: 139–148. doi:10.1016/j.cell.2015.10.048.Microbiota-modulated

21. Cluzeau T, McGraw KL, Irvine B, Masala E, Ades L, Basiorka AA, et al. Pro-inflammatory proteins S100A9 and tumor necrosis factor-α suppress erythropoietin elaboration in myelodysplastic syndromes. Haematologica. 2017;102: 2015–2020. doi:10.3324/haematol.2016.158857

22. Flint HJ, Duncan SH, Louis P. The impact of nutrition on intestinal bacterial communities. Current Opinion in Microbiology. 2017;38: 59–65. doi:10.1016/j.mib.2017.04.005

23. Morrison DJ, Preston T. Gut Microbes Formation of short chain fatty acids by the gut microbiota and their impact on human metabolism. Gut microbes. 2016;7: 189–200. doi:10.1080/19490976.2015.1134082

24. Crost EH, Le Gall G, Laverde-Gomez JA, Mukhopadhya I, Flint HJ, Juge N. Mechanistic insights into the cross-feeding of Ruminococcus gnavus and Ruminococcus bromii on host and dietary carbohydrates. Frontiers in Microbiology. 2018;9: 1–13. doi:10.3389/fmicb.2018.02558

25. Severyn CJ, Brewster R, Andermann TM. Microbiota modification in hematology: Still at the bench or ready for the bedside? Blood Advances. 2019;3: 3461–3472. doi:10.1182/bloodadvances.2019000365

26. Zeevi D, Korem T, Zmora N, Israeli D, Rothschild D, Weinberger A, et al. Personalized Nutrition by Prediction of Glycemic Responses. Cell. 2015;163: 1079– 1094. doi:10.1016/j.cell.2015.11.001

27. Zaki H, Boyd KL, Kastan MB, Lamkanfi M, Kanneganti D. Integrity and Mortality During Experimental Colitis. 2010;32: 379–391. doi:10.1016/j.immuni.2010.03.003.The

28. Meers S, Kasran A, Boon L, Lemmens J, Ravoet C, Boogaerts M, et al. Monocytes are activated in patients with myelodysplastic syndromes and can contribute to bone marrow failure through CD40–CD40L interactions with T helper cells. Leukemia. 2007;21: 2411–2419. doi:10.1038/sj.leu.2404940

29. Qin H, Wilson CA, Lee SJ, Zhao X, Benveniste EN. LPS induces CD40 gene expression through the activation of NF-kappaB and STAT-1alpha in macrophages and microglia. Blood. 2005/07/14. 2005;106: 3114–3122. doi:10.1182/blood-2005-02-0759

30. Barreyro L, Chlon TM, Starczynowski DT. Chronic immune response dysregulation in MDS pathogenesis. Blood. 2018;132: 1553–1560. doi:10.1182/blood-2018-03-784116

31. de Sousa JC, da Nóbrega Ito M, Costa MB, Farias IR, de Paula Borges D, de Oliveira RT, et al. Dysregulation of interferon regulatory genes reinforces the concept of chronic immune response in myelodysplastic syndrome pathogenesis. Hematological Oncology. 2019;37: 523–526. doi:10.1002/hon.2608

32. Yang Y, Wang H, Kouadir M, Song H, Shi F. Recent advances in the mechanisms of NLRP3 inflammasome activation and its inhibitors. Cell Death and Disease. 2019;10. doi:10.1038/s41419-019-1413-8

33. Derrien M, Belzer C, de Vos WM. Akkermansia muciniphila and its role in regulating host functions. Microbial Pathogenesis. 2017;106: 171–181. doi:10.1016/j.micpath.2016.02.005

34. Routy B, Le Chatelier E, Derosa L, Duong CPM, Alou MT, Daillère R, et al. Gut microbiome influences efficacy of PD-1-based immunotherapy against epithelial tumors. 2018.

